# Detection of SARS-CoV-2 by Real-Time PCR under challenging pre-analytical conditions reveals independence of swab media and cooling chain

**DOI:** 10.1101/2020.07.21.20158154

**Authors:** Sabrina Summer, Ralf Schmidt, Anna Nele Herdina, Isabella Krickl, Julia Madner, Georg Greiner, Florian J. Mayer, Nicole Perkmann-Nagele, Robert Strassl

## Abstract

With global demand for SARS-CoV-2 testing ever rising, shortages in commercially available viral transport media pose a serious problem for laboratories and health care providers. For reliable diagnosis of SARS-CoV-2 and other respiratory viruses, executed by Real-time PCR, the quality of respiratory specimens, predominantly determined by transport and storage conditions, is crucial. Therefore, our aim was to explore the reliability of minimal transport media, comprising saline and the CDC recommended Viral Transport Media (HBSS VTM), for the diagnosis of SARS-CoV-2 and other respiratory viruses (influenza A, respiratory syncytial virus, adenovirus, rhinovirus and human metapneumovirus) compared to commercial products, such as the Universal Transport Media (UTM). We question the assumptions, that the choice of medium and temperature for storage and transport affect the accuracy of viral detection by RT-PCR. Both alternatives to the commercial transport medium (UTM), namely the CDC viral transport media (HBSS VTM) and saline, allow adequate detection of SARS-CoV-2 and other respiratory viruses, regardless of the storage temperature and time.

Our study revealed the high resilience of SARS-CoV-2 and other respiratory viruses, enabling proper detection in clinical specimens even after long-time storage at high temperatures, independent of the transport medium’s composition.

## INTRODUCTION

The rapid spread of the novel coronavirus SARS-CoV-2, probably of zoonotic origin ^[1]^, has led to a continuing COVID-19 pandemic ^[2]^. Extensive laboratory testing for SARS-CoV-2 is currently among the most effective measures to curtail the spread of COVID-19, together with quarantine and social distancing measures ^[3,4]^. Real-time PCR (RT-PCR), typically performed from upper respiratory specimens, represents the current gold standard for SARS-CoV-2 detection. The World Health Organization (WHO) recommends cooled storage (2-8°C) and the transport of respiratory specimens in specific viral transport medium (VTM) up to 5 days ^[3]^. Viral transport medium exists in several formulas, all consisting of a buffered salt solution, a complex source of protein and/or amino acids, and antimicrobial agents ^[5]^. As an alternative to the commercially available products, the CDC provided a standard operating procedure for the production of viral transport medium suitable for viral detection ^[6]^.

The COVID-19 pandemic and the associated demand of mass testing have severely challenged worldwide supplies of commercial viral transport media as well as commercial swab kits and reagents for SARS-CoV-2 RT-PCR ^[5,6]^. Due to the increased demand and the shortage in supply, the compatibility of alternative minimal transport buffers, such as 0.9% saline solution, with viral detection in diagnostics have to be assessed. The primary aim of this study was to define time and temperature-dependent alterations detecting SARS-CoV-2 RNA in clinical samples stored in commercially available universal transport media (UTM), in the CDC-provided alternative HBSS VTM ^[6]^ and in 0.9% NaCl at 4 to 28 °C. Additionally, this study aimed to provide evidence on storage and transport conditions of other respiratory viruses, including influenza A, respiratory syncytial virus, adenovirus, rhinovirus and human metapneumovirus, for future recommendations.

## RESULTS

### HBSS VTM and saline are adequate alternatives to UTM as transport media for SARS-CoV-2 specimens in diagnostics

To obtain data on the thermal long-time stability of SARS-CoV-2 RNA for diagnostics assessed by RT-PCR, swab specimens of 15 positively-tested SARS-CoV-2 patients were stored in the viral transport medium prepared by the authors according to the CDC recommendation (HBSS VTM) and saline (0.9% NaCl), respectively, over 28 days at three different temperatures (4 °C, 21 °C and 28 °C). Additionally, 12 swab specimens, 6 stored in the Universal Transport Medium from Cepheid, Copan Diagnostics (UTM) and 6 in saline, were stored at 21°C for 14 days for comparing the alternative media, HBSS VTM and saline, to commercial UTM. The original Cq-values detected at time point zero (T0) for all samples were below 45 cycles. For each transport medium, we analyzed a range of SARS-CoV-2 ++ (Cq <27, *n*=22, mean Cq 22.16 ± 0.62) to SARS-CoV-2 + (Cq ≥27, *n*=12, mean Cq 29.86 ± 0.48) specimens. Besides, we also included 3 negative specimens as control.

A comparison of the changes in the Cq-values of specimens transported in VTM and saline over 28 days did not show any significant differences between the two transport media (Figure 1 and Supplementary Dataset). Besides, no significant differences between SARS-CoV-2 specimens stored in HBSS VTM and UTM at 21 °C were observed either (mean ΔCq E T14 for UTM −0.3583 ± 0.3683, HBSS VTM 0.6344 ± 0.1524, NaCl 0.6753 ± 0.2770, *p*=0.101, mean ΔCq S T14 for UTM 1.365 ± 0.6909, HBSS VTM 1.716 ± 0.2671, NaCl 1.867 ± 0.3079, *p*=0.637; Figure 2). In specimens stored in VTM (HBSS VTM and UTM) an increased tendency for bacterial contamination could be observed by the macroscopic formation of white sediment and drop in pH; however, this has not impaired the analysis. In general, the virus could be consistently detected in both transport media under different storage conditions over 4 weeks (Figure 1 and Supplementary Dataset). However, further analysis suggests slight differences in the stability of the target genes to diagnose SARS-CoV-2 (S and E gene). Both genes seem to be on-trend more stable at 4 °C and 28 °C compared to 21 °C (Figure 1, Supplementary Figure S1 and Supplementary Dataset); notably, no significant changes between the different transport media could be observed. At 21 °C (RT) the S gene differs in its stability according to the storage medium; the ΔCq-values of the samples stored in VTM (HBSS VTM and UTM) and saline, respectively, vary after 14 and up to 28 days; some specimens show an increased drop in Cq-values at 28 days (Supplementary Figure S1 and Supplementary Dataset). On the contrary, the variations in the ΔCq-values of the E gene were more prominent between the two transport media at 21°C at the 28-day time point. Specimens stored in saline at RT showed on trend an increase in the ΔCq after 7 days of storage (Figure 1 and Supplementary Dataset).

**Figure 1:**
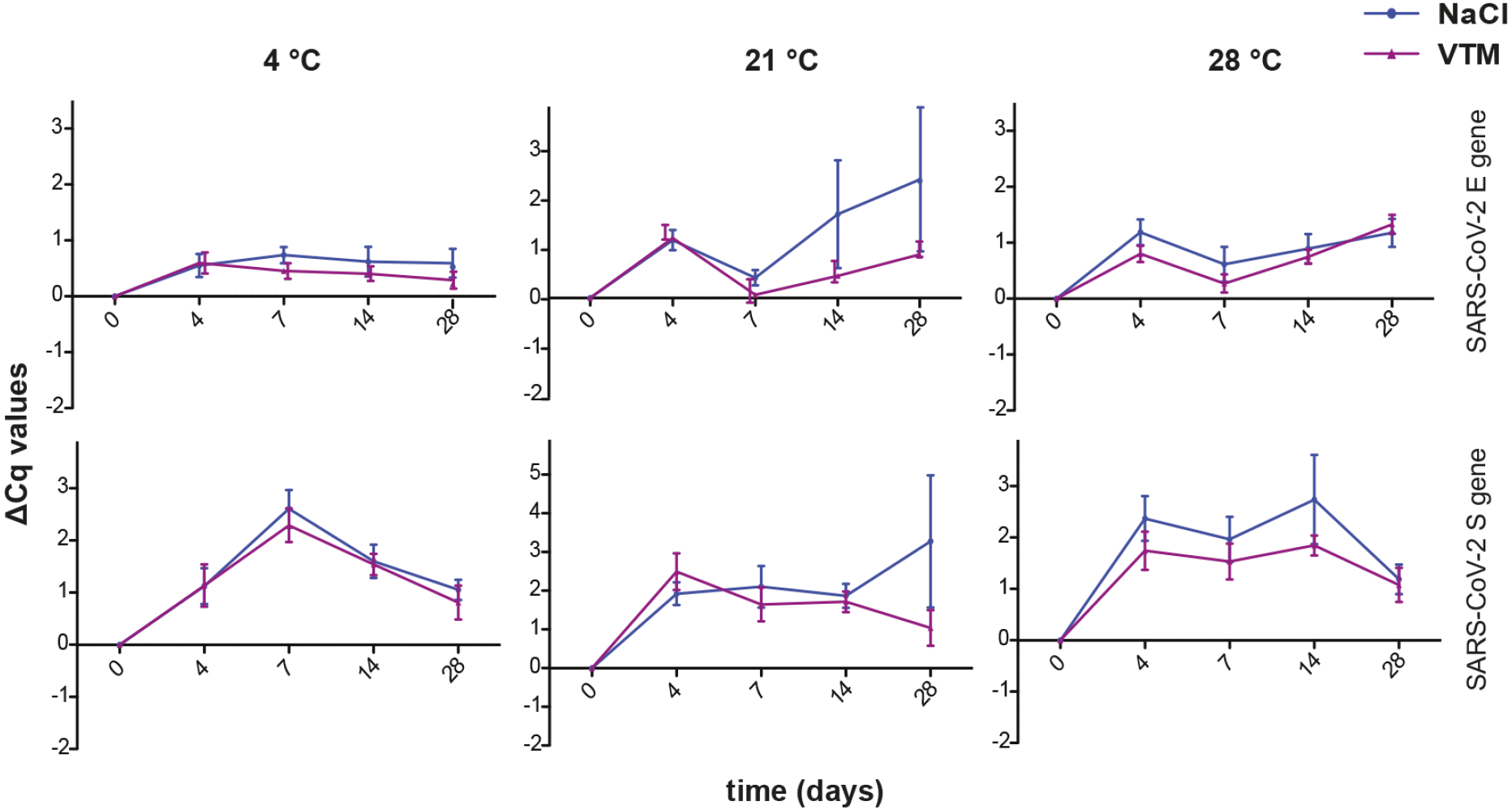
VTM and saline are both suitable transport media for SARS-CoV-2 specimens at high storage temperatures. RT-PCR measurements of clinical specimens positive for SARS-CoV-2 stored in NaCl and VTM medium over 4 weeks at 3 different temperatures (4 °C, 21 °C and 28 °C). ΔCq-values ± SEM for SARS-CoV-2 at 4 °C and 28 °C *n* = 12, SARS-CoV-2 at 21 °C *n* = 22 are shown. * *p*-values<0.05, repeated measurement ANOVA.

**Figure 2:**
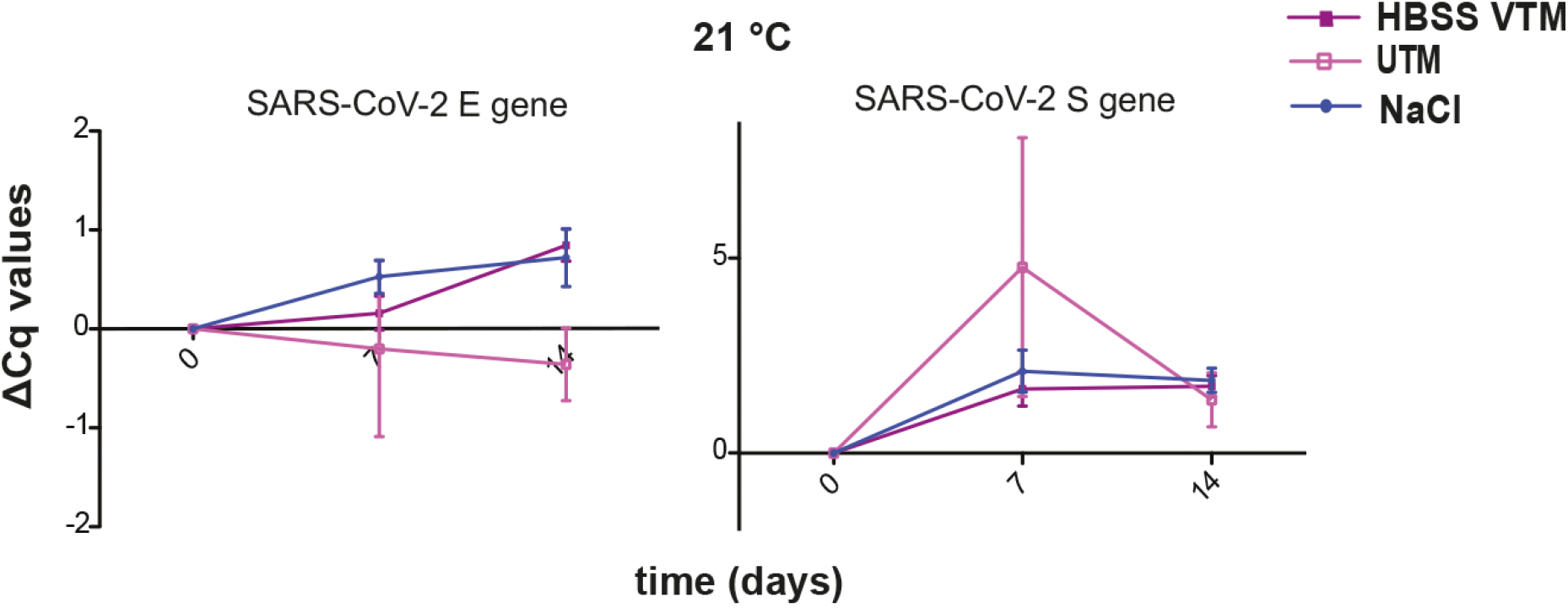
HBSS VTM and saline are both suitable alternatives to UTM as transport media for SARS-CoV-2 specimens at 21 °C. RT-PCR measurements of clinical specimens positive for SARS-CoV-2 stored in NaCl, HBSS VTM and UTM medium over 14 days at 21 °C. ΔCq-values ± SEM for NaCl and HBSS VTM *n* = 16 and UTM *n* = 6 are shown. * *p*-values<0.05, repeated measurement ANOVA. Comparison of SARS-CoV-2 + and SARS-CoV-2 ++ samples have not revealed any significant differences in general (Figure 3). In both groups the viral RNA levels were adequately stable for RT-PCR detection, independent of the storage conditions. However, the stability of SARS-CoV-2 + samples kept at 4 °C and 28 °C varied significantly from SARS-CoV-2 ++ (* 4 °C *p* E=0.0008, *p* S=<0.0001; 28 °C *p* E=0.0049, *p* S=0.0001), however, no significant changes in the Cq-values could be observed for the specimens stored at 21 °C (*p* E=0.73, *p* S=0.60).

**Figure 3:**
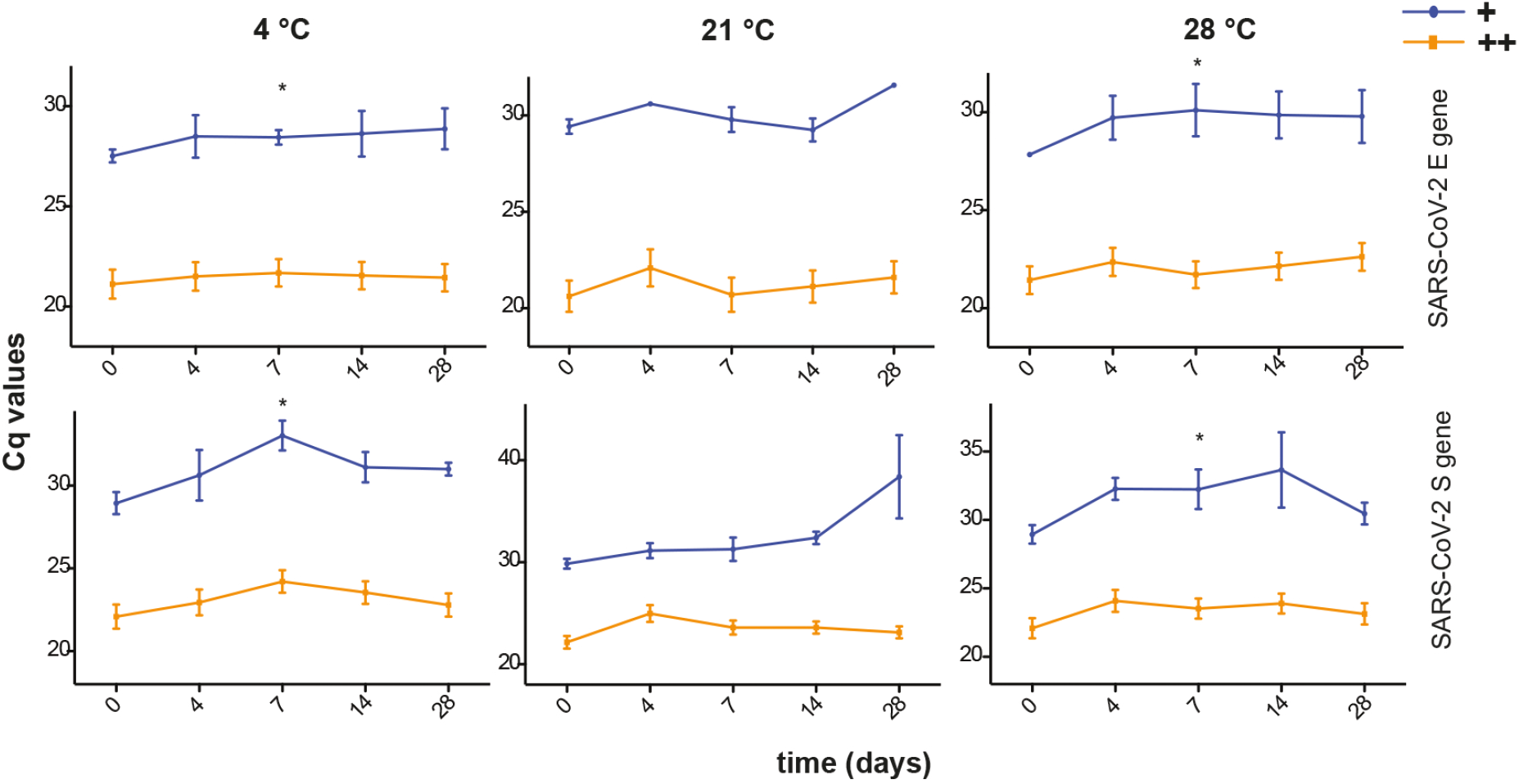
Long-term storage, even at high temperatures, allows adequate detection of weakly positive SARS-CoV-2 specimens. RT-PCR measurements of clinical specimens strongly and weakly positive for SARS-CoV-2 stored in NaCl and VTM medium, respectively, over 4 weeks at 3 different temperatures (4 °C, 21 °C and 28°C). Cq-values ± SEM for SARS-CoV-2 + at 4 °C *n* = 3-4; at 21 °C *n* = 10-12 and 28 °C *n* = 2-4 and for SARS-CoV-2 ++ at 4 °C *n* = 20; at 21 °C *n* = 17-22 and 28 °C *n* = 20-22 are shown. * *p*-values<0.05, repeated measurement ANOVA and Wilcoxon signed rank test.

### High stability of other respiratory viruses in VTM and saline at higher temperatures

To evaluate the thermal stability of other respiratory viruses for their detection by RT-PCR, three swabs collected from patients positively tested for influenza A (mean Cq 30.34 ± 0.70), respiratory syncytial virus (mean Cq 23.49 ± 0.53), adenovirus (mean Cq 26.13 ± 1.10), rhinovirus (mean Cq 23.56 ± 0.40) and human metapneumovirus (mean Cq 24.94 ± 0.79), respectively, stored in either viral transport medium (HBSS VTM or UTM) or saline over 4 weeks at constant 4 °C, 21 °C and 28 °C, respectively, (Figure 4) were compared. Detection of the various respiratory viruses was stable, even at higher temperatures over the observation time; however, influenza A, respiratory syncytial virus and rhinovirus stored at 21 °C showed a destabilization of the detected target gene, mainly when stored in saline (Figure 4, Supplementary Figure S2 and Supplementary Dataset). In general, Cq-values of influenza A, respiratory syncytial virus, adenovirus, rhinovirus and human metapneumovirus stored in VTM and saline were consistently detectable and showed no significant decrease in Cq-values over time (Supplementary Dataset).

**Figure 4:**
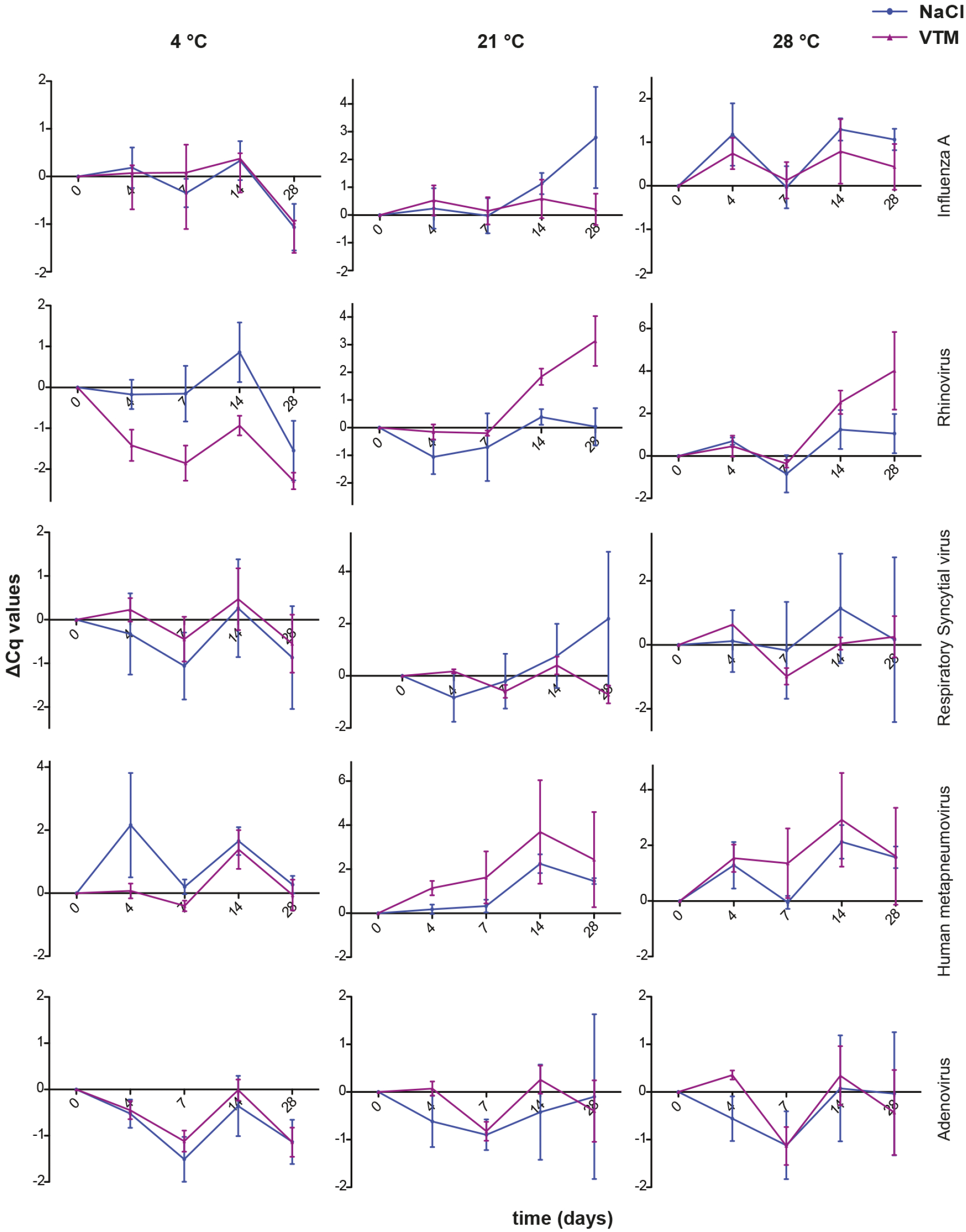
VTM and saline are both suitable transport media for respiratory virus specimens at high storage temperatures. RT-PCR measurements of clinical specimens positive for influenza A, respiratory syncytial virus, rhinovirus, human metapneumovirus and adenovirus stored in NaCl and VTM medium over 4 weeks at 3 different temperatures (4 °C, 21 °C and 28 °C). *n* = 3 are shown. * *p*-values<0.05, repeated measurement ANOVA.

## DISCUSSION

The COVID-19 pandemic has re-emphasized the importance of molecular diagnostics for the acquisition of timely results aiding in the containment of the pathogen. In the case of SARS-CoV-2, previous studies have shown that the virus, in contrast to other respiratory viruses, like influenza, is not susceptible to heat, supporting the assumption that the prevalence of SARS-CoV-2 will not be seasonal ^[7,8]^. Future demand for mass testing, requiring complex logistics with long transports, required a study determining the thermostability of the virus over a longer time period. High outdoor temperatures or storage at non-cooled areas combined with a long transport time of respiratory specimens could affect the virus detection capabilities of molecular methods such as RT-PCR. This could especially be a problem for mass testing and testing in countries with poor infrastructure where rapid, cooled transport and the availability of commercial transport media for respiratory specimens cannot be guaranteed. In the present study, we evaluated the stability of SARS-CoV-2 and a variety of other respiratory viruses, representing different families varying in size, structure and genetic material, for diagnostic analysis by RT-PCR at temperatures up to 28 °C, considering possible effects mediated by the transport medium. In general, it is assumed that the sensitivity of viruses to heat increases with size and the existence of an envelope; hence, non-enveloped viruses are the most stable. However, there are only a few studies reporting thermostability of structurally different respiratory viruses under identical conditions ^[9,10]^. In our study, we could not detect a correlation between structure and stability. Comparing the enveloped SARS-CoV-2 virus with non-enveloped respiratory viruses (rhinovirus and adenovirus), we detected no significant differences in the long-term thermostability. The RNA of all viral species observed in this study showed a high thermal stability. Using positively tested clinical specimens, we demonstrated the remarkable stability of the viral RNA upon long-time storage (up to 28 days) at high temperatures (21 and 28 °C), independent of the transport medium (Figure 1 and Supplementary Figure S1). Storage in alternatives to the commercially available UTM medium, such as the CDC recommended viral transport medium (HBSS VTM) and saline, were equally suitable for adequate detection of the two target genes E (betacoronavirus-specific) and S (SARS-CoV-2-specific) in specimens characterized by a low and high viral load, respectively (Figure 3). However, at higher temperatures, samples stored in VTM (HBSS VTM and UTM) proved to be more susceptible to bacterial contamination, even when supplemented with antibiotics. Nevertheless, bacterial contamination has neither affected sample preparation nor target detection.

The relatively high resilience towards higher temperatures has been previously reported for some respiratory viruses. Our observations confirmed this for a variety of respiratory viruses, including SARS-CoV-2 ^[11-15]^, however, our study presented additional data on “time” as an essential factor potentially affecting viral detection. For the handling of SARS-CoV-2 and other respiratory virus specimens in diagnostics, the WHO recommends a maximal storage of 5 days at 2-8 °C ^3^. Our findings that even a minimal buffered system such as saline provides a suitable viral transport medium for stable long-time storage of the clinical samples, lacking significant changes in the detection levels of the target genes, not even in case of higher temperatures, could be advantageous to increase future testing capacities. In regards to the minimal alterations in the detected RNA signal, which are not significant, SARS-CoV-2 is highly resilient to high temperatures. Its detection by RT-PCR is sufficiently robust for the diagnosis of COVID-19, even in seasons or countries characterized by higher outdoor temperatures.

In summary, we have shown that the stability of SARS-CoV-2 RNA in human specimens seems to robust, even compared to other coronaviruses (Figure 1 and Supplementary Figure S1) ^[16]^. Increasing temperatures and long-term storage conditions did not affect its stability. Besides, our data confirmed that even saline and the CDC recommended transport media (HBSS VTM), which are cheaper and more easily accessible, provide adequate alternatives to the commercially available UTM. Both can be deployed for transporting, and even for long-term preservation of SARS-CoV-2 specimens at higher outdoor temperatures or non-cooled transport vehicles. Our data on the thermal stability of respiratory viruses from different families will help assessing the likelihood of viral stability through long, non-cooled transports or sample storages and it emphasizes that an increased capacity of testing for widespread screening of SARS-CoV-2 and early diagnosis of COVID-19 can be achieved.

## METHODS

### Sample collection

This study was performed at the Medical University of Vienna, Department of Laboratory Medicine, Division of Clinical Virology during the pandemic of SARS-CoV-2 in 2020. During this period, nasopharyngeal swabs were collected from patients showing acute respiratory tract infections. The archived respiratory samples were stored in either saline (0.9% NaCl) or UTM (Universal Transport Medium, Cepheid, Copan Diagnostics) and frozen at −80°C after the initial patient diagnosis. Stored respiratory specimens from the archive of the Division of Clinical Virology at the Medical University of Vienna, which have been tested positive by routine PCR within the last 2 years for other respiratory viruses (influenza A, respiratory syncytial virus, adenovirus, rhinovirus and human metapneumovirus), were used for the evaluation.

### Sample preparation

After one freeze-thaw cycle 12 anonymized SARS-CoV-2-positive samples and 3 anonymized SARS-CoV-2-negative respiratory swab samples were diluted 50-fold in 3.5 ml self-made viral transport medium (HBSS VTM, HBSS buffer based recipe after CDC recommendations: 500 ml HANKS Balanced Salt Solution HBSS (Gibco), 2% FBS (Gibco), 100 µg/ml Gentamycin (B. Braun Melsungen AG), 0.5 µg/ml Amphotericin B (Cheplapharm)) ^6^ and saline (0.9% NaCl; B. Braun Melsungen AG), respectively, for each temperature; independent of the medium they were archived in. 700 µl of the pool were used for RNA extraction at the certain time point. Following the initial measurement at time point zero (T0), the samples were stored at different temperatures (4 °C, 21 °C and 28 °C) for up to 28 days. Total RNA was extracted after 4 (T4), 7 (T7), 14 (T14) and 28 (T28) days. pH values were determined, which served as a surrogate parameter for bacterial contamination.

For comparison of HBSS VTM and saline, respectively, with UTM, 12 anonymized, undiluted SARS-CoV-2-positive samples from 12 individual patients, 6 collected in UTM (Universal Transport Medium, Cepheid, Copan Diagnostics) and 6 in saline, were stored at 21 °C after the initial measurement (T0). RNA was extracted after 7 (T7) and 14 (T14) days.

### RNA isolation and RT-PCR analysis

Total RNA of SARS-CoV-2 and the other respiratory viruses was extracted from 500 µl of the clinical samples by the AltoStar Automation System AM16 (Altona Diagnostics, Hamburg, Germany) according to the manufacturer’s instructions and eluted in 45 µl elution buffer (Altona Diagnostics, Hamburg, Germany). RT-PCR was carried out using the RealStar SARS-CoV-2 RT-PCR kit (Altona Diagnostics, Hamburg, Germany) targeting the SARS-CoV-2-specific S gene and the betacoronavirus-specific E gene on a Bio-Rad CFX Connect Real-Time PCR Detection System (Bio-Rad Laboratories, Hercules, California, USA). Cq-values (cycle threshold values as calculated specifically by the Bio-Rad CFX Manager software) less than 45 cycles for both tested targets were interpreted as positive for SARS-CoV-2. Positive specimens were further divided into SARS-CoV-2 ++ (Cq <27) and SARS-CoV-2 + (Cq ≥27). Internal controls were used to confirm measurement integrity.

The presence of other respiratory viruses used in this study, influenza virus A, adenovirus, respiratory syncytial virus, rhinovirus and human metapneumovirus, were confirmed by TaqMan-based RT-PCR using the LightCycler Multiplex RNA Virus Master (Roche Diagnostics, Rotkreuz, Switzerland) on a Bio-Rad CFX Connect Real-Time PCR Detection System (Bio-Rad Laboratories, Hercules, California, USA). RT-PCR was performed using the routine testing setup for respiratory viruses at the Medical University of Vienna, Department of Laboratory Medicine, Division of Clinical Virology by the following cycling protocol: 8 minutes at 53 °C, 30 seconds 95 °C, followed by 50 cycles of 1 second at 95 °C, 40 seconds at 60 °C, 1 second at 72 °C and 30 seconds at 40 °C. Primers and probes ^[17-21]^ are listed in Table 1.

**Table 1:**
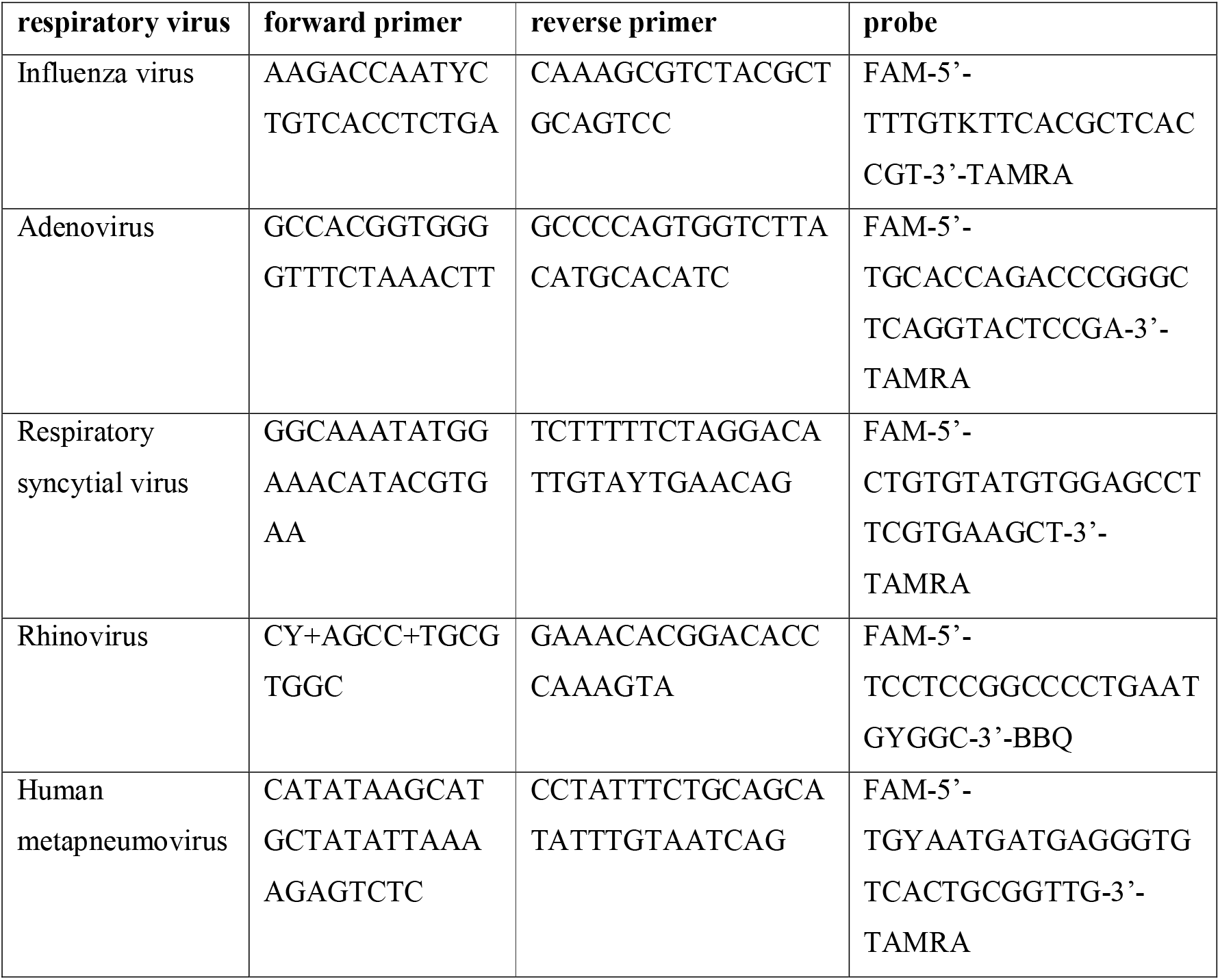
RT-PCR primers and probes used in this study.

### Data analysis

Cq-values were exported from CFX Manager Dx Software version 3.1. ΔCq-values were calculated in Microsoft Excel Version 2019 as the differences between the target Cq measured at the dedicated day and temperature and the target Cq of the initial measurement (T0). “Not detectable” signals (N/A) were set to 45 cycles for analysis (total number of cycles of the RT-PCR).

Details on sample sizes are provided in the figure legends. Statistical analysis and calculating the mean Cq-values were done in GraphPad Prism Version 5.0 (GraphPad). Kolmogorov-Smirnov normality test was done in GraphPad Prism Version 5.0 (GraphPad). In most cases, a two-tailed repeated measurement ANOVA was performed with adjustments for multiple comparisons, following the Bonferroni post-hoc test when indicated. In one case, when data was not normally distributed, a Wilcoxon signed-rank test was done. Significant differences *(p*<0.05) were marked by a * above the corresponding curves in the figures.

## Data Availability

Anonymized data will be made available by the authors upon reasonable request.

## STATEMENT OF ETHICAL APPROVAL

The local Institutional Review Board deemed the study exempt from review.

Respiratory swab samples from patients with suspected SARS-CoV-2 infection were first assessed at the Division of Clinical Virology at the Department of Laboratory Medicine at the Medical University of Vienna. A collection of left-over material of positive samples was anonymized and then randomized for purposes of this study before stability analysis. Thus, in correspondence with representatives of the ethical board of the Medical University of Vienna, no separate consultation of the ethics committee was necessary to guarantee ethical standards for conducting biomedical research as required by the Declaration of Helsinki.

## DATA AVAILABILITY

The datasets generated and analyzed during the current study are available from the authors upon reasonable request.

## ACKNOWLEDGEMENTS

We thank Karin Mildner for excellent technical assistance in RT-PCR detection of other respiratory viruses.

## AUTHOR CONTRIBUTIONS STATEMENT

R.S., R.S.*, G.G., N.PN. and F.J.M. first planned the study, all authors refined the concept. S.S. and AN.H. conducted the experiments, with technical assistance by I.K. and J.M..

S.S. and R.S. analyzed the data and designed the figures. S.S. and AN.H. wrote the manuscript and all authors revised it and approved the final version. All authors have accepted responsibility for the entire content of this manuscript and approved its submission.

## ADDITIONAL INFORMATION

The author(s) declare no competing interests. No external funding was received for this study.

