## Supplementary Table and Figures for "Detection of SARS-CoV-2 by Real-Time PCR under challenging pre-analytical conditions reveals independence of swab media and cooling chain"

### Supplementary Figures

Supplementary Figure S1

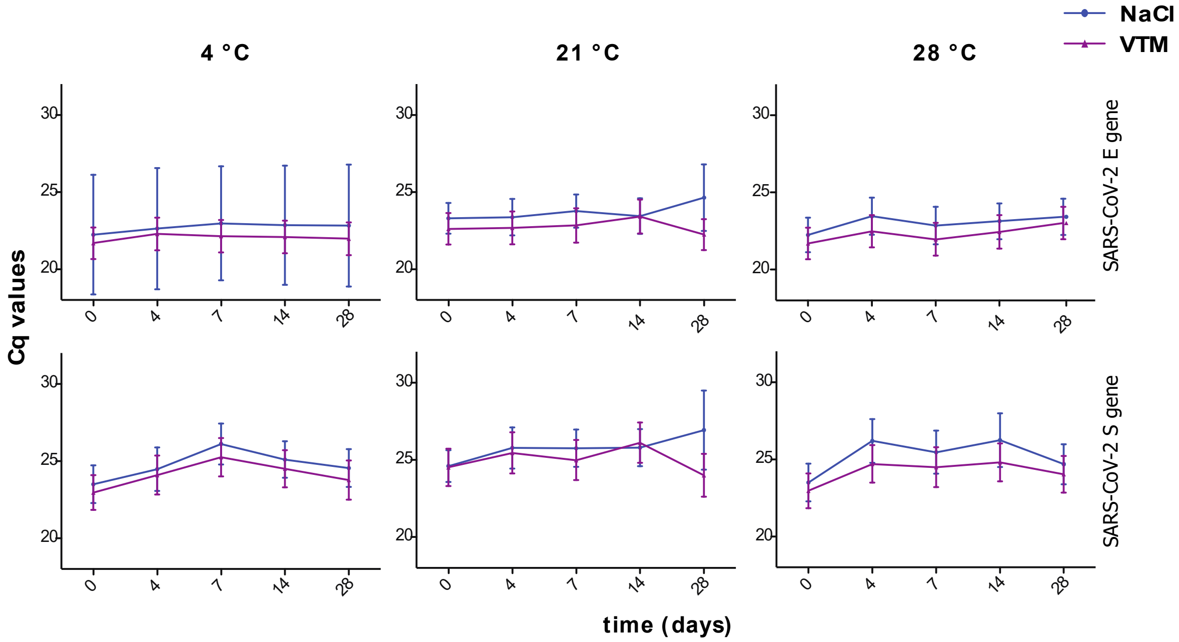


**Supplementary Figure S1: VTM and saline are both suitable transport media for SARS-CoV-2 specimens at high storage temperatures**

RT-PCR measurements of clinical specimens positive for SARS-CoV-2 and other respiratory viruses (Influenza A, respiratory syncytial virus, rhinovirus, human metapneumovirus and adenovirus) stored in NaCl and HBSS VTM medium over 28 days at three different temperatures (4 °C, 21 °C and 28 °C). Cq-values ± SEM for SARS-CoV-2 at 4 °C and 28 °C *n* = 11-12, SARS-CoV-2 at 21 °C *n* = 22 and the other respiratory viruses *n* = 3 are shown. * *p*-values<0.05 for differences between media, repeated measurement ANOVA.

Supplementary Figure S2

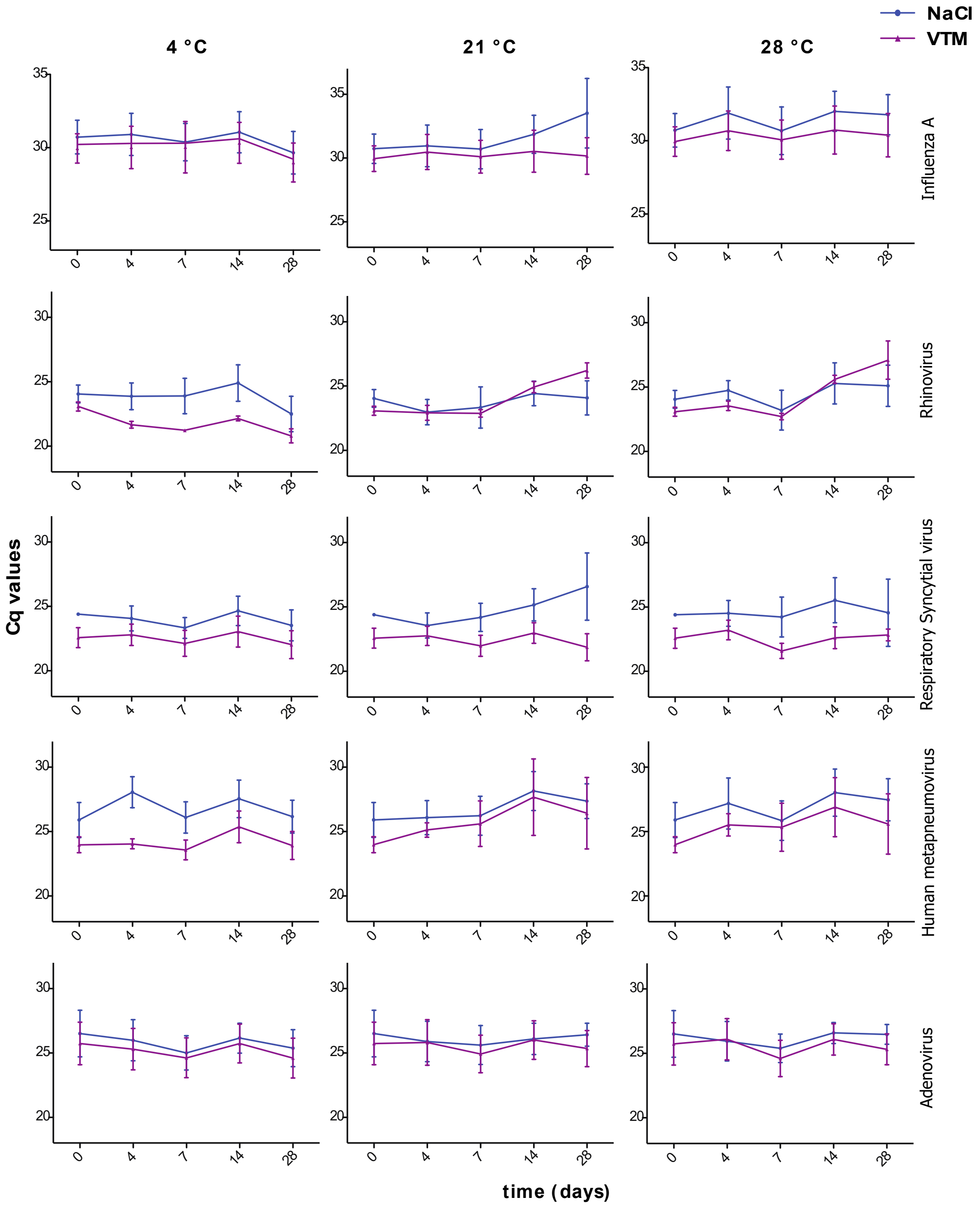


**Supplementary Figure S2: VTM and saline are both suitable transport media for respiratory virus specimens at high storage temperatures**

RT-PCR measurements of clinical specimens positive for influenza A, respiratory syncytial virus, rhinovirus, human metapneumovirus and adenovirus stored in NaCl and VTM medium over 4 weeks at 3 different temperatures (4 °C, 21 °C and 28 °C). n = 3 are shown. * p-values<0.05, repeated measurement ANOVA.
